# Plans to vaccinate children for COVID-19: a survey of US parents

**DOI:** 10.1101/2021.05.12.21256874

**Authors:** Chloe A. Teasdale, Luisa N. Borrell, Spencer Kimball, Michael L. Rinke, Madhura Rane, Sasha A. Fleary, Denis Nash

## Abstract

In a national online survey of 2,074 US parents conducted in March 2021, 49.4% reported plans to vaccinate their child for COVID-19 when available. Lower income and less education were associated with greater parental vaccine hesitancy/resistance, while safety, effectiveness and lack of need were the primary reasons for vaccine hesitancy/resistance.

## INTRODUCTION

As of April 2021, more than 3.7 million US children have been diagnosed with COVID-19 and almost 300 have died.^1^ Although children are less likely to experience severe disease and mortality from COVID-19 infection compared to adults, younger children, those with underlying health conditions and from communities disproportionately affected by the COVID-19 epidemic are vulnerable to severe disease.^2-6^ In addition, COVID-19 infection can lead to multi□inflammatory syndrome in children (MIS□C) and there is some evidence that children may experience long COVID with ongoing symptoms, including fatigue and pain.^7-9^ Preventing COVID-19 in children is of critical importance and, to date, mask wearing and social distancing, including closing or reducing time at school, have been the primary approaches for infection control.^10,11^

While no COVID-19 vaccines are yet approved for children <16 years of age, clinical trials in pediatric populations are underway.^12^ Once vaccines are available for use in children, parental willingness to vaccinate will be critical for protecting children from COVID-19 infection, reducing risk in schools, increasing population immunity levels and ending the epidemic.^13-15^ A safe and effective vaccine will allow children to return to normal activities, and help relieve the social isolation as well as other negative impacts the epidemic has had on children’s mental health and well-being.^16,17^

It is estimated that more than two thirds of the US population will need to be vaccinated in order to end sustained transmission of COVID-19.^18,19^ In a survey of US adults conducted in March 2021, more than 60% had either received vaccination or were planning to receive it and according to the Centers for Disease Control and Prevention’s COVID Data Tracker, by the end of April 2021, approximately 147 million or 44.4% of adults in the US had received at least one dose of an approved COVID-19 vaccine.^20,21^ There is limited evidence on the extent to which parents intend to vaccinate their children for COVID-19 when a pediatric vaccine is available. We report findings from a national survey measuring plans among US parents to vaccinate their children against COVID-19, reasons for parents not wanting to vaccinate children and the relationship between a parent’s own vaccination status and their plans to vaccinate their child.

## METHODS

### Study Design

We conducted a community-based, non-probability survey of English and Spanish speaking US parents and caregivers (‘parents’) of children ≤12 years of age using a panel recruited online by Qualtrics, an online platform that sources participants through social media and partner networks. Adults ≥18 years who identified as primary caregivers of a child ≤12 years who had taken a child for ≥1 medical visit in the past 2 years were eligible (n=2,074). Data were collected from March 9 through April 2, 2021. Survey weights were based on race, ethnicity, sex, education and region and designed to provide national estimates of US parents of children ≤12 years according to 2019 US Census data.^22^ Ethics approval was received from the CUNY School of Public Health and Health Policy institutional review board.

The study outcome was the proportion of parents reporting that they want to vaccinate the youngest child in the household. Participants were asked “when a vaccine to prevent COVID-19 is approved for children, would you want your child to receive the vaccine”, with response options “yes”, “no” and “unsure”. Those responding “no” or “unsure” were asked “why do you not want your child to receive the COVID-19 vaccine?” and could choose multiple options. In addition to reporting demographics and household data, parents reported whether they had received or plan to get the COVID-19 vaccine themselves with responses options including already received COVID-19 vaccination, plan to receive when available, unsure, will not get the vaccine and prefer not to answer.

### Data Analyses

Descriptive statistics (unweighted counts and weighted percentages) on the total sample population and prevalence estimates are presented according to vaccination plans (Yes, No, Unsure). Rao adjusted Pearson chi-squared tests were used to compare parental vaccination plans according to sample characteristics. Poisson regression models with robust standard errors were fitted to estimate prevalence ratios (PR) and their confidence intervals (CI) comparing parents planning to vaccinate to those responding “no” or “unsure” (combined) and were adjusted for demographic and household characteristics. All analyses were adjusted for survey weights to provide national estimates. Parental vaccination status was examined by grouping together those responding they had already received with those who planned to receive vaccination and those responding that they were unsure or preferred not to answer (parents who reported they would not get vaccinated comprised their own group). We examined the association between parental vaccine status with reported intentions to vaccinate children using Rao adjusted Pearson chi-squared tests to compare proportions. Analyses were conducted in SAS 9.4 (SAS Institute Inc., Cary, NC, USA).

## RESULTS

Among 2,074 US parents surveyed, 49.4% said they intended to vaccinate the youngest child in their household (median child age: 4.8 years; interquartile range: 4.5-5.1) for COVID-19 when a pediatric vaccine is approved, 25.6% said they would not and 25.0% said they were unsure (Table 1). Among parents responding that they would not or were unsure whether they would vaccinate their child, 78.2% reported potential safety or effectiveness concerns, 23.0% reported that they did not believe children need to be vaccinated, and 8.5% and 11.2% reported religious or medical reasons, respectively.

**Table 1.** **Estimated prevalence of parental plans to vaccinate children ≤12 years for COVID-19 by parent and child characteristics –** United States, March 9-April 2, 2021

|  |  |  | When a COVID-19 vaccine available for children, will you want your child to be vaccinated |  |  |  |  |  |  |
| --- | --- | --- | --- | --- | --- | --- | --- | --- | --- |
|  | Sample |  | Yes |  | No |  | Unsure |  |  |
| Characteristics | N | %* | %^ | 95% CI | % | 95% CI | % | 95% CI | p-value† |
| Total sample | 2074 | 100.0 | 49.4 | 46.9-52.0 | 25.6 | 23.3-27.8 | 25.0 | 22.7-27.3 |  |
| **Child age, median in years (IQR) 4.8 (4.4-5.1)** |  |  |  |  |  |  |  |  |  |
| <2 years | 371 | 18.6 | 37.2 | 31.2-43.1 | 34.1 | 28.4-39.8 | 28.7 | 23.3-34.1 | <0.0001 |
| 2-6 years | 831 | 40.2 | 48.0 | 43.9-52.1 | 25.4 | 21.8-29.0 | 26.6 | 22.8-30.4 |  |
| 7-12 years | 872 | 14.1 | 56.5 | 52.7-60.3 | 21.8 | 18.7-25.0 | 21.7 | 18.5-24.9 |  |
| **Child sex** |  |  |  |  |  |  |  |  |  |
| Male | 1022 | 50.3 | 50.4 | 46.8-54.1 | 26.3 | 23.0-29.5 | 23.3 | 20.2-26.4 | 0.50 |
| Female | 1046 | 49.5 | 48.6 | 45.0-52.1 | 24.7 | 21.7-27.8 | 26.7 | 23.3-30.0 |  |
| Missing§ | 6 | 0.3 | -- | -- | -- | -- | -- | -- |  |
| **Child race/ethnicity** |  |  |  |  |  |  |  |  |  |
| Black non-Hispanic | 200 | 10.6 | 41.1 | 33.6-48.6 | 30.5 | 23.4-38.7 | 28.3 | 21.3-35.3 | <0.0001 |
| Asian | 99 | 3.7 | 77.5 | 68.1-86.9 | 4.8 | 0.7-8.9 | 17.7 | 8.8-26.5 |  |
| White non-Hispanic | 1099 | 50.6 | 52.1 | 48.6-55.6 | 24.4 | 21.4-27.4 | 23.5 | 20.3-26.7 |  |
| Hispanic | 488 | 25.9 | 46.2 | 40.8-51.6 | 27.5 | 22.7-32.3 | 26.3 | 21.5-31.0 |  |
| Other non-Hispanic | 188 | 9.3 | 42.3 | 33.8-50.9 | 29.3 | 21.8-36.7 | 28.4 | 21.4-35.4 |  |
| **Parent age** |  |  |  |  |  |  |  |  |  |
| 18-29 years | 366 | 20.3 | 33.5 | 27.5-39.5 | 39.1 | 32.2-45.0 | 27.4 | 22.0-32.7 | <0.0001 |
| 30-44 years | 1387 | 65.1 | 53.4 | 50.3-56.5 | 23.2 | 20.6-25.9 | 23.4 | 20.6-26.2 |  |
| 45-64 years | 321 | 14.6 | 54.2 | 47.8-60.6 | 17.1 | 12.1-22.1 | 28.7 | 22.9-34.5 |  |
| **Parent sex** |  |  |  |  |  |  |  |  |  |
| Male | 794 | 39.3 | 67.9 | 64.0-71.8 | 18.0 | 14.9-21.2 | 14.1 | 11.0-17.1 | <0.0001 |
| Female | 1270 | 60.1 | 37.4 | 34.2-40.5 | 30.4 | 27.4-33.4 | 32.2 | 29.1-35.3 |  |
| Transgender/other§ | 10 | 0.6 | -- | -- | -- | -- | -- | -- |  |
| **Parent race/ethnicity** |  |  |  |  |  |  |  |  |  |
| Black non-Hispanic | 219 | 11.3 | 40.5 | 33.6-47.4 | 29.7 | 23.1-36.2 | 29.8 | 23.4-36.3 | <0.0001 |
| Asian | 129 | 4.6 | 71.9 | 63.2-80.7 | 7.8 | 3.0-12.5 | 20.3 | 12.3-28.3 |  |
| White non-Hispanic | 1159 | 53.3 | 52.1 | 48.7-55.5 | 24.3 | 21.4-27.2 | 23.6 | 20.6-26.7 |  |
| Hispanic | 467 | 25.4 | 48.0 | 42.5-53.6 | 28.3 | 23.3-33.3 | 23.7 | 19.1-28.4 |  |
| Other non-Hispanic | 100 | 5.3 | 29.8 | 19.9-38.6 | 31.8 | 21.1-42.5 | 38.4 | 26.4-50.3 |  |
| **Child has health insurance** |  |  |  |  |  |  |  |  |  |
| Yes | 1914 | 92.0 | 48.9 | 46.2-51.5 | 25.5 | 23.2-27.8 | 25.6 | 23.3-28.0 | 0.26 |
| No | 149 | 7.3 | 56.0 | 46.1-66.0 | 27.4 | 18.5-36.3 | 16.6 | 8.5-24.6 |  |
| Don't know§ | 11 | 0.6 | -- | -- | -- | -- | -- | -- |  |

| Child attending in person school/daycare $\geq 1$ day per week | | | | | | | | | |
| --- | --- | --- | --- | --- | --- | --- | --- | --- | --- |
| Yes | 1098 | 49.7 | 56.3 | 52.9-59.7 | 23.0 | 20.1-25.9 | 20.7 | 17.9-23.5 | <0.0001 |
| No | 969 | 50.0 | 42.6 | 38.9-46.4 | 28.2 | 24.8-31.6 | 29.1 | 25.6-32.7 |  |
| Don't know§ | 7 | 0.2 | -- | -- | -- | -- | -- | -- |  |
| Number of children $\leq 12$ years in household | | | | | | | | | |
| 1 | 1059 | 50.8 | 50.9 | 47.4-54.4 | 24.2 | 21.3-27.2 | 24.9 | 21.8-28.0 | 0.02 |
| 2 | 751 | 34.7 | 51.9 | 47.6-56.1 | 25.4 | 21.6-29.2 | 22.7 | 19.1-26.3 |  |
| 3 or more | 264 | 14.4 | 38.6 | 31.3-45.9 | 30.7 | 24.0-37.4 | 30.7 | 23.3-38.0 |  |
| Parent education (highest completed) |  |  |  |  |  |  |  |  |  |
| High school or less | 482 | 30.7 | 35.3 | 29.7-40.9 | 32.2 | 27.1-37.3 | 32.5 | 27.2-37.9 | <0.0001 |
| Some college | 546 | 31.4 | 43.1 | 38.7-47.4 | 28.5 | 24.6-32.4 | 28.4 | 24.5-32.3 |  |
| Completed college or more | 1015 | 36.5 | 66.0 | 62.8-69.2 | 18.5 | 15.8-21.1 | 15.6 | 13.1-18.0 |  |
| Missing§ | 31 | 1.4 | -- | -- | -- | -- | -- | -- |  |
| Household income, USD |  |  |  |  |  |  |  |  |  |
| <\$25,000 | 331 | 20.3 | 35.3 | 29.0-41.7 | 33.3 | 27.5-39.2 | 31.4 | 25.3-37.5 | <0.0001 |
| \$25,000-\$49,999 | 472 | 24.6 | 37.6 | 32.4-42.7 | 31.1 | 26.3-35.8 | 31.4 | 26.5-36.3 | |
| \$50,000-\$99,999 | 587 | 27.4 | 54.2 | 49.6-58.7 | 23.8 | 19.8-27.8 | 22.0 | 18.1-25.9 | |
| ≥\$100,000 | 617 | 23.9 | 72.2 | 68.0-76.7 | 15.2 | 11.7-18.6 | 12.6 | 9.7-15.6 | |
| Missing | 67 | 3.8 | 25.3 | 12.4-38.1 | 26.6 | 12.1-41.1 | 48.1 | 32.7-63.6 |  |
| US Region |  |  |  |  |  |  |  |  |  |
| Northeast | 550 | 15.7 | 56.6 | 51.9-61.4 | 20.6 | 16.3-24.9 | 22.8 | 18.9-26.6 | <0.0001 |
| South | 684 | 39.0 | 44.3 | 40.0-48.6 | 28.6 | 24.6-32.5 | 27.1 | 23.1-31.1 |  |
| Midwest | 442 | 21.0 | 44.0 | 38.9-49.0 | 31.3 | 26.5-36.2 | 24.7 | 20.1-29.3 |  |
| West | 398 | 24.3 | 57.8 | 52.3-63.4 | 19.0 | 14.8-23.2 | 23.2 | 18.3-28.0 |  |

Abbreviations: CI = confidence interval; IQR = interquartile range; USD = US dollars.
\*Survey weights applied to sample to represent US population by race, ethnicity, sex, education and region.
^Weighted percents are prevalence estimates of US parents reporting they were planned to vaccinate their youngest child, were not willing or were not sure.
† p-values from Rao adjusted Pearson Chi-squared tests to compare expected to observed frequencies among groups by characteristic for parent's willingness to vaccinate their youngest child (i.e. whether willing to vaccinate youngest child differed by sex of the child, etc.).
§Categories are not presented in the table as they yielded unreliable standard error estimates.

In adjusted models Asian parents were 38% more likely to report intentions to vaccinate their children compared to non-Hispanic Whites (adjusted PR [aPR]: 1.38;95% CI: 1.19-1.60) (Table 2). Parents less likely to report plans to vaccinate their children were female (aPR: 0.69;95%CI: 0.62-0.77), had high school education or less (aPR: 0.73;95%CI: 0.62-0.86) and had household income <$25,000 (0.75;95%CI: 0.64-0.88) (Table 2).

**Table 2.**
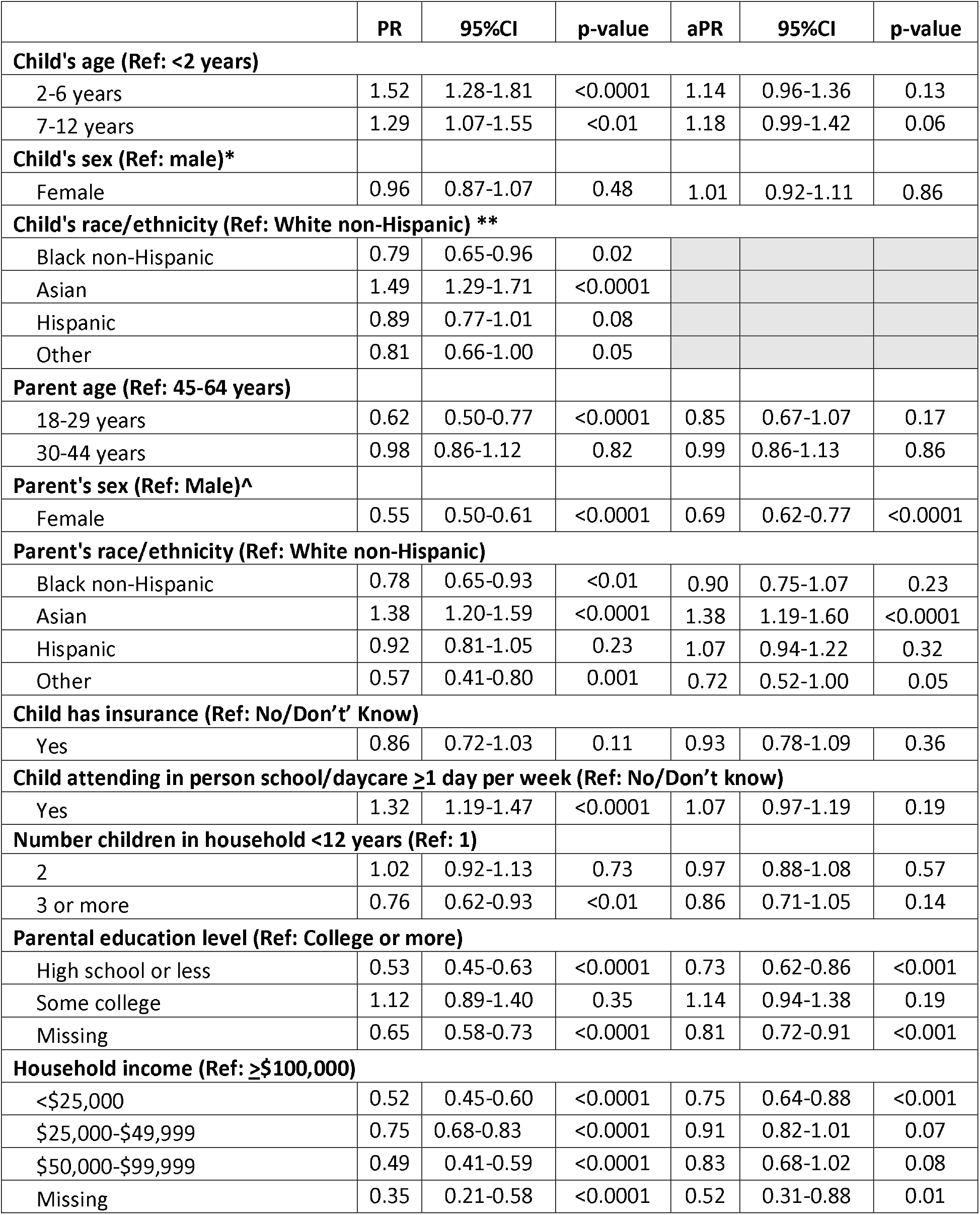
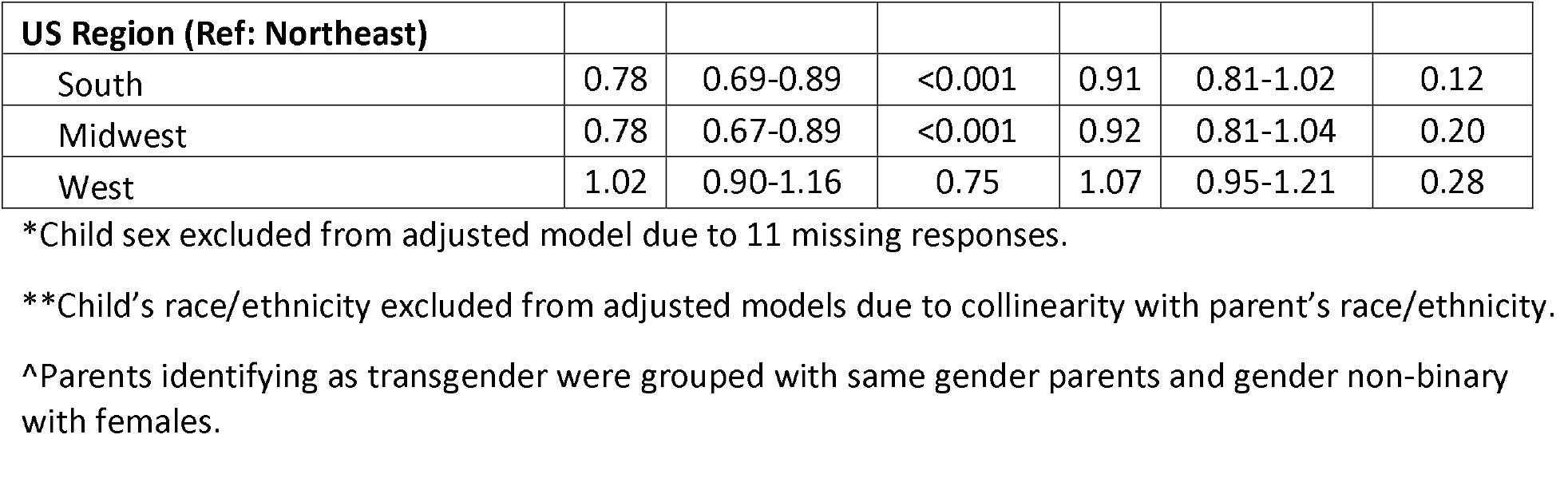
**Characteristics associated with parental plans to vaccinate children ≤12 years for COVID-19 –** United States, March 9-April 2, 2021

Across all parents surveyed, 16.1% reported having received COVID-19 vaccination, 34.2% planned to receive it when available to them, 24.6% were unsure whether they would get vaccinated, 22.4% reported planning not to get vaccinated and 2.6% preferred not to answer. Among parents reporting that they had gotten or would get vaccinated, 85.2% said they would get their child vaccinated for COVID-19, 10.0% said they were unsure and 4.8% reported they would not vaccinate their child. Parents who were unsure or did not plan to vaccinate themselves were much less likely to report wanting to vaccinate their children: 19.5% of parents who were unsure about getting vaccinated themselves said they would vaccinate their child, 60.4% were unsure about vaccinating a child and 20.1% said they would not. Only 5.7% of parents who said they would not get vaccinated themselves reported planning to vaccinate their child, 15.6% were unsure and 78.8% said they would not vaccinate their child (p<0.0001) (Figure 1).

**Figure 1.**
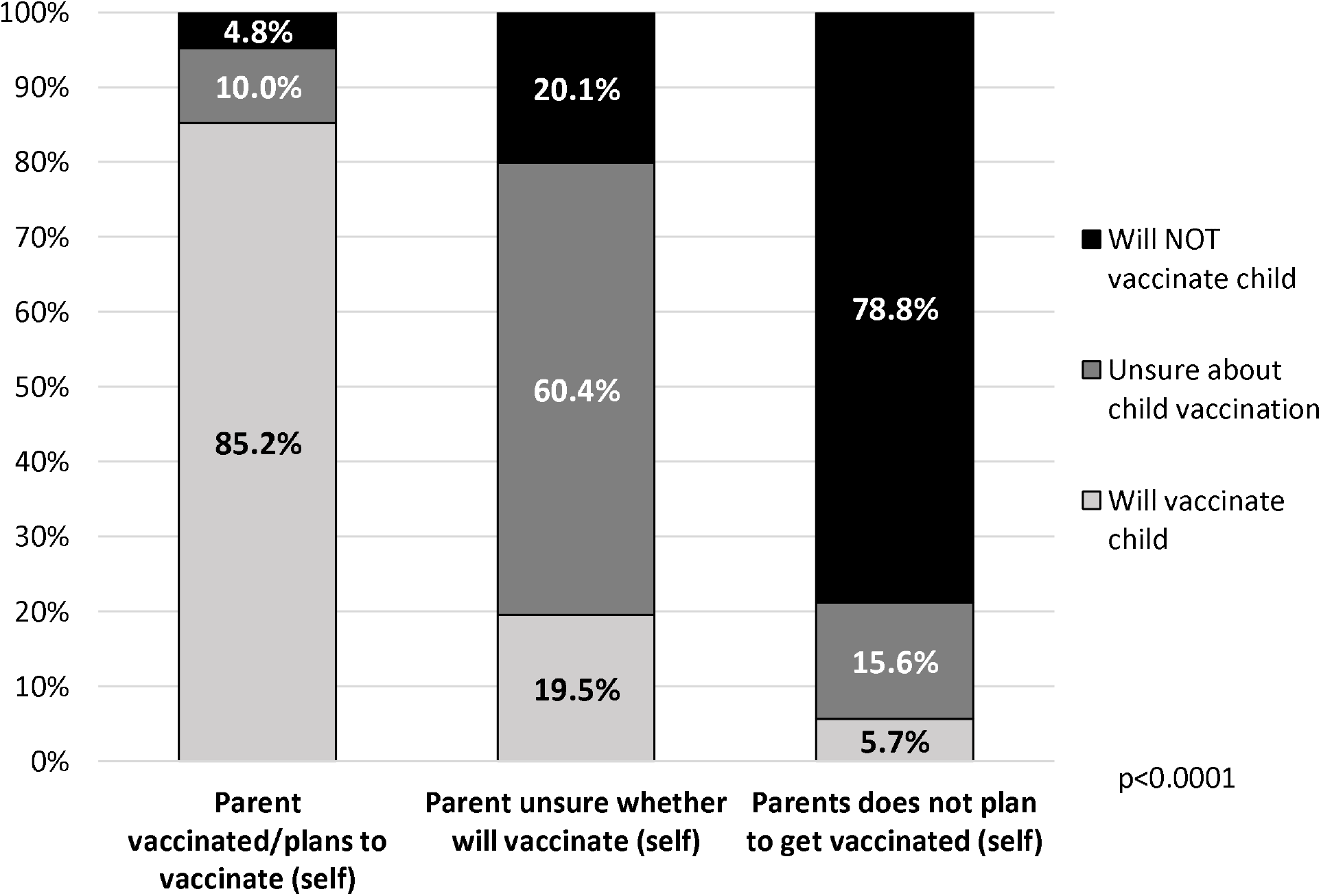
**Parental intentions to vaccinate children against COVID-19 according to parents’ own vaccination status –** United States, March 9-April 2, 2021

## DISCUSSION

As of March 2021, only half of US parents reported plans to have their youngest child receive a COVID-19 vaccine when they become available for pediatric populations. The primary concerns reported by parents were safety and effectiveness, as well as perceived lack of need. Female parents, those with less education and lower income were least likely to report plans to vaccinate their children. We observed a strong correlation between a parent’s own vaccination status or hesitancy with hesitancy to vaccinate their child when a pediatric vaccine is available. These findings have important implication for pediatric vaccine policies and roll-out planning over the coming months.

Our findings showing only half of parents reporting they will vaccinate their child, and 26% saying they will not, are concerning given the estimated vaccination coverage required to reach herd immunity.^18,19^ There are few data with which to compare our findings regarding intentions among US parents to vaccinate their children for COVID-19. Therefore, our findings provide important information with regard to the current state of willingness, as well as the characteristics of parents who are hesitant. We found that, similar to previous studies of other childhood vaccinations, lower parental education was a predictor of vaccine hesitancy.^23,24^ Our findings also suggest that while Asian parents report being more likely to report wanting to vaccinate their child, female parents and those with lower education were less likely to want their child to receive COVID-19 vaccines.^25^ These findings provide critical information that can be used to inform strategies to increase awareness about COVID-19 vaccination for children to increase uptake when available.

The majority of parents surveyed who reported they will not or are unsure whether they will vaccinate their child cited safety and effectiveness as the primary reason (78%) and almost a quarter reported that children are at low risk for COVID-19 infection and do not need to be vaccinated. These findings are consistent with a previous report of vaccine hesitancy for routine childhood immunization and influenza where safety and lack of perceived susceptibility are the main reasons for delaying or not accepting vaccination.^26^ Our findings suggest that providing evidence of the safety of COVID-19 vaccination will be important for increasing uptake, as well as educating parents about the importance of vaccination for children.

Approximately 50% of parents in our survey reported that they already had or planned to get vaccinated for COVID-19 which is somewhat lower than a poll conducted among all US adults by Kaiser News Network (also conducted in March 2021) which included more older adults compared to our sample.^20^ Studies in US adults have also shown more vaccine hesitancy among females which was consistent with our findings that female parents were less likely to report wanting to vaccinate their child.^25^ We also found a strong correlation between parental vaccination status/hesitancy and plans for vaccinating the child. These findings could suggest that an additional benefit of increasing adult vaccination coverage may be increased uptake for children; however, this requires further study.

There are several limitations to our analysis. Our survey focused on children 12 years of age and younger to collect information about younger children but we therefore do not have data on adolescents. The survey data are self-reported and thus subject to recall, response and social desirability bias. In addition, while our survey was weighted to reflect the US population of parents based on 2019 census estimates, it was conducted online therefore excluded parents without access to the internet and thus may not accurately reflect the US population by income and educational attainment. Finally, our survey was conducted prior to the pause of the Johnson & Johnson vaccine distribution due to safety concerns which may be contributing to increased vaccine hesitancy.^27^

Overall, our findings suggest that targeted efforts will be needed to ensure high uptake and coverage of COVID-19 vaccination in US children.^14^ They also show that providing evidence of the safety of vaccines and educating parents about the importance of vaccinating children may help in efforts to reach the levels of coverage that will be needed to reach herd immunity.

## Data Availability

study data are available upon request

## Abbreviations

(COVID-19): Coronavirus disease 2019
(US): United States
(PR): Prevalence ratio
(aPR): adjusted prevalence ratio
(CI): confidence interval

## Notes

### Competing Interest Statement

The authors have declared no competing interest.

### Funding Statement

This project was funded by the Institute for Implementation Science in Population Health (ISPH) of the City University of New York (CUNY) Graduate School of Public Health and Health Policy (SPH).

### Author Declarations

Ethics approval was received from the CUNY School of Public Health and Health Policy institutional review board.

## References

1. Amercian Academy of Pediatrics (AAP) and Children’s Hospital Association. Children and COVID-19: State Data Report Version: April 29, 2021. Available from: https://services.aap.org/en/pages/2019-novel-coronavirus-covid-19-infections/children-and-covid-19-state-level-data-report/. Accessed

2. Stower H. Clinical and epidemiological characteristics of children with COVID-19. Nat Med. 2020;26(4):465.

3. Kim L, Whitaker M, O’Halloran A, et al. Hospitalization Rates and Characteristics of Children Aged <18 Years Hospitalized with Laboratory-Confirmed COVID-19 - COVID-NET, 14 States, March 1-July 25, 2020. MMWR Morb Mortal Wkly Rep. 2020;69:1081–1088.

4. Williams N, Radia T, Harman K, Agrawal P, Cook J, Gupta A. COVID-19 Severe acute respiratory syndrome coronavirus 2 (SARS-CoV-2) infection in children and adolescents: a systematic review of critically unwell children and the association with underlying comorbidities. Eur J Pediatr. 2021;180:689–697.

5. Felsenstein S, Hedrich CM. SARS-CoV-2 infections in children and young people. Clin Immunol. 2020;220:108588.

6. Oualha M, Bendavid M, Berteloot L, et al. Severe and fatal forms of COVID-19 in children. Arch Pediatr. 2020;27:235–238.

7. Kest H, Kaushik A, DeBruin W, Colletti M, Goldberg D. Multisystem Inflammatory Syndrome in Children (MIS-C) Associated with 2019 Novel Coronavirus (SARS-CoV-2) Infection. Case Rep Pediatr. 2020:8875987.

8. Buonsenso D, Munblit D, De Rose C, et al. Preliminary evidence on long COVID in children. Acta Paediatr. 2021. doi: 10.1111/apa.15870.

9. Felsenstein S, Willis E, Lythgoe H, et al. Presentation, Treatment Response and Short-Term Outcomes in Paediatric Multisystem Inflammatory Syndrome Temporally Associated with SARS-CoV-2 (PIMS-TS). J Clin Med. 2020;9:3293.

10. Auger KA, Shah SS, Richardson T, et al. Association Between Statewide School Closure and COVID-19 Incidence and Mortality in the US. JAMA. 2020;324:859–870.

11. Centers for Disease Control and Prevention (CDC). Help Stop the Spread of COVID-19 in Children. 2021. Avaiable from: https://www.cdc.gov/coronavirus/2019-ncov/daily-life-coping/children/protect-children.html. Accessed May 3, 2021.

12. Centers for Disease Control and Prevention (CDC). Summary Document for Interim Clinical Considerations for use of COVID-19 Vaccines Currently Authorized in the United States. Available from: https://www.cdc.gov/vaccines/covid-19/downloads/summary-interim-clinical-considerations.pdf. Accessed April 28, 2021.

13. Rhodes ME, Sundstrom B, Ritter E, McKeever BW, McKeever R. Preparing for A COVID-19 Vaccine: A Mixed Methods Study of Vaccine Hesitant Parents. J Health Commun. 2020;25:831–837.

14. Bubar KM, Reinholt K, Kissler SM, et al. Model-informed COVID-19 vaccine prioritization strategies by age and serostatus. Science. 2021;371:916–921.

15. Velavan TP, Pollard AJ, Kremsner PG. Herd immunity and vaccination of children for COVID-19. Int J Infect Dis. 2020;98:14–15.

16. Liu JJ, Bao Y, Huang X, Shi J, Lu L. Mental health considerations for children quarantined because of COVID-19. Lancet Child Adolesc Health. 2020;4:347–349.

17. Fegert JM, Vitiello B, Plener PL, Clemens V. Challenges and burden of the Coronavirus 2019 (COVID-19) pandemic for child and adolescent mental health: a narrative review to highlight clinical and research needs in the acute phase and the long return to normality. Child Adolesc Psychiatry Ment Health. 2020;14:20.

18. Randolph HE, Barreiro LB. Herd Immunity: Understanding COVID-19. Immunity. 2020;52:737–741.

19. Kwok KO, Lai F, Wei WI, Wong SYS, Tang JWT. Herd immunity - estimating the level required to halt the COVID-19 epidemics in affected countries. J Infect. 2020;80:e32–e33.

20. Kaiser Health News (KHN). Covid Vaccine Hesitancy Drops Among All Americans, New Survey Shows. KFF COVID-19 Vaccine Monitor 2021. Available from: https://khn.org/news/article/covid-vaccine-hesitancy-drops-among-americans-new-kff-survey-shows/. Accessed: May 3, 2021.

21. Centers for Disease Control and Prevention (CDC). COVID Data Tracker: COVID-19 Vaccinations in the United States. 2021; https://covid.cdc.gov/covid-data-tracker/#vaccinations. Accessed May 3, 2021.

22. US Census Bureau (USC). Census QuickFacts (2019). Available from: https://www.census.gov/quickfacts/fact/table/US/PST045219. Accessed January 5, 2021.

23. Kempe A, Saville AW, Albertin C, et al. Parental Hesitancy About Routine Childhood and Influenza Vaccinations: A National Survey. Pediatrics. 2020;146:e20193852..

24. Santibanez TA, Nguyen KH, Greby SM, et al. Parental Vaccine Hesitancy and Childhood Influenza Vaccination. Pediatrics. 2020;146:e2020007609.

25. Khubchandani J, Sharma S, Price JH, Wiblishauser MJ, Sharma M, Webb FJ. COVID-19 Vaccination Hesitancy in the United States: A Rapid National Assessment. Journal Community Health. 2021;46:270–277.

26. Siddiqui M, Salmon DA, Omer SB. Epidemiology of vaccine hesitancy in the United States. Hum Vaccin Immunother. 2013;9:2643–2648.

27. Frellick M. J&J pause magnifies worry about COVID-19 vaccine hesitancy. Medscape, Arpil 13, 2021. Available from: https://www.medscape.com/viewarticle/949236. Accessed: May 3, 2021.

